# Methylation-derived Inflammatory Measures and Lung Cancer Risk and Survival

**DOI:** 10.1101/2021.05.24.21257709

**Authors:** Naisi Zhao, Mengyuan Ruan, Devin C. Koestler, Jiayun Lu, Karl T. Kelsey, Elizabeth A. Platz, Dominique S. Michaud

**Affiliations:** Department of Public Health & Community Medicine, Tufts University School of Medicine, Tufts University, Boston, MA; Department of Biostatistics & Data Science, University of Kansas Medical Center, Kansas City, KS; University of Kansas Cancer Center, Kansas City, KS; Department of Epidemiology, Johns Hopkins Bloomberg School of Public Health, Baltimore, MD; The Sidney Kimmel Comprehensive Cancer Center at Johns Hopkins, Baltimore, MD; Department of Epidemiology, Brown University, Providence, RI; Department of Pathology and Laboratory Medicine, Brown University, Providence, RI

**Keywords:** Lung cancer, DNA methylation, Methylation-based inflammation measures, C-reactive Protein, mdNLR

## Abstract

**Background:** Examining inflammation-related DNA methylation alterations in blood could help elucidate the role of inflammation in lung cancer etiology and aid discovery of factors that are key to lung cancer development and progression. In a nested case-control study, we estimated the neutrophil-to-lymphocyte ratio using a validated index, methylation-derived NLR (mdNLR), and quantified DNA methylation levels at loci previously linked with circulating concentrations of C-reactive protein (CRP). We examined associations between these measures and lung cancer risk, and among the cases, lung cancer survival, using pre-diagnostic blood samples of cases (median of 14 years before diagnosis) and controls in the CLUE I/II cohorts. Our analyses controlled for self-reported smoking and methylation-predicted cumulative smoking in order to better focus our examinations on the DNA methylation marks that are informative of the immune response profile.

**Results:** Using conditional logistic regression and further adjusting for BMI, batch effects, and a smoking-based methylation score, we observed a 47% increased risk of non-small cell lung cancer (NSCLC) for one standard deviation increase in mdNLR (n = 150 pairs; OR: 1.47 [1.08, 2.02]) and found the estimated CRP Scores to be inversely associated with risk of NSCLC risk after additionally adjusting for methylation-predicted pack-years (n = 150 pairs; Score 1 OR: 0.57 [0.40, 0.81]; Score 2 OR: 0.62 [0.45, 0.84]; Score 3 OR: 0.65 [0.44, 0.95]). Using Cox proportional-hazards models and adjusting age, sex, smoking status, methylation-predicted pack-years, BMI, batch effect, and stage, we observed a 27% increased risk of dying from lung cancer for one standard deviation increase in mdNLR (n = 145 deaths in 205 cases; HR: 1.27 [1.08, 1.50]). A 50% increased risk of dying from lung cancer for one standard deviation increase in mdNLR was observed for NSCLC cases (n = 103 deaths in 149 cases; HR: 1.50 [1.19, 1.89]).

**Conclusions:** A better understanding of inflammation-associated methylation-based biomarkers in lung cancer development could provide insight into critical pathways that may help identify new markers of early disease and survival.

## Background

Lung cancer is the leading cause of cancer deaths in the US, projected to account for 21.7% of all cancer-deaths in 2021 (1). A large percentage of lung cancer patients are diagnosed at an advanced stage (2) and five-year relative survival rates for those patients are between 3-6% (3). Thus, early detection remains a key strategy to improve survival. However, the currently recommended strategy for lung cancer screening – low-dose computed tomography (LDCT) for persons 50 to 80 years old with a least a 20 pack-year smoking history and currently smoke or have quit within the past 15 years – is expensive and has a high false positive rate (4). Modifying current lung cancer screening strategy by performing risk stratification could help prioritize LDCT screening and optimize secondary prevention. We propose that immune system markers could be incorporated into such risk stratification tools to help identify persons at higher risk of lung cancer to target for screening.

While smoking is the most important risk factor for lung cancer in the population, there is growing evidence that the immune system, in response to or independent of smoking, plays an important role in lung cancer development, acting potentially through the genesis of chronic inflammation (5). Furthermore, it is plausible that inflammatory profiles prior to lung cancer diagnosis are associated with lung cancer-specific survival. Markers of systemic inflammation, including elevated levels of C-reactive protein (CRP) and the peripheral blood neutrophil to lymphocyte ratio (NLR), also have been identified as robust marker of cancer associated inflammation (6, 7). Elevated CRP levels (6), elevated serum levels of pro-inflammatory cytokines (8-10), increased neutrophil counts and decreased lymphocyte counts(11, 12),and polymorphisms in inflammation-related genes (13-16) have been associated with increased lung cancer risk. These inflammatory measures have also been associated with poor survival of lung cancer patients in several retrospective and a few prospective studies (17-19). In addition, both experimental and epidemiologic studies support a role for chronic inflammation as a hallmark of cancer development and progression (6, 20-23).

Understanding the role of inflammation in lung cancer etiology could aid discovery of factors that stimulate, suppress, or modulate cancer risk and cancer survival. We posit that such understanding could be gained by examining DNA methylation alterations in blood that are associated with the systemic immune response. In addition, measuring methylation-derived inflammatory responses using pre-diagnostic samples provides the opportunity to capture informative individual systemic inflammatory profiles years prior to diagnosis, potentially shedding light on risk factors key to lung cancer development and progression, e.g., underlying genetics, exposure to environmental risk factors, and behavior risk factors.

In the current study, we predicted peripheral blood leukocyte composition and a neutrophil to lymphocyte index using validated DNA methylation markers (mdNLR) and quantified DNA methylation levels at loci previously linked with circulating concentrations of CRP, and evaluated their associations with lung cancer risk and lung cancer-specific survival. To address this question, we used pre-diagnostic blood samples of cases and controls obtained from the CLUE I/II cohorts. Our analyses controlled for self-reported smoking and methylation-predicted cumulative smoking in order to better focus our examinations on the DNA methylation marks that are informative of the immune response profile (24).

## Results

### Population characteristics

Characteristics of the 208 lung cancer cases and their 208 matched controls included in this analysis are presented in Table 1. Over 99% of the majority of participants were white. The median time between blood draw and lung cancer diagnosis was 14 years. The median age at blood draw in 1989 was 59 and 57 years in cases and controls, respectively. Overall, 55% of cases and controls were women and 11% were never smokers (Table 1).

**Table 1.** Baseline characteristics of lung cancer cases and matched controls, CLUE I/II

|  | Controls | Cases |  |  |
| --- | --- | --- | --- | --- |
|  |  | All lung cancers | Non-small cell lung cancer | Small-cell lung cancer |
| N | 208 | 208 | 150 | 29 |
| Median Age (Range, Years) | 56 [28, 81] | 59 [30, 83] | 58 [36, 83] | 53 [30, 71] |
| Median Time before diagnosis (Range, years) |  | 14 [0, 29] | 14 [0, 28] | 12 [2, 29] |
| Sex |  |  |  |  |
| Male | 95 (45.7%) | 95 (45.7%) | 61 (40.7%) | 16 (55.2%) |
| Female | 113 (54.3%) | 113 (54.3%) | 89 (59.3%) | 13 (44.8%) |
| Race |  |  |  |  |
| White | 208 (100%) | 205 (98.6%) | 149 (99.3%) | 28 (96.6%) |
| Cigarette Smoking Status |  |  |  |  |
| Never Smoker | 22 (10.6%) | 22 (10.6%) | 15 (10.0%) | 1 (3.4%) |
| Ever Smoker | 80 (38.5%) | 80 (38.5%) | 56 (37.3%) | 10 (34.5%) |
| Current Smoker | 106 (51.0%) | 106 (51.0%) | 79 (52.7%) | 18 (62.1%) |
| Median Smoking Intensity (Range, Cig/Day) | 20 [0, 80] | 20 [0, 80] | 20 [0, 70] | 20 [0, 80] |
| Cigar or Pipe Smoking |  |  |  |  |
| Never | 176 (84.6%) | 178 (85.6%) | 132 (88.0%) | 25 (86.2%) |
| Ever | 28 (13.5%) | 26 (12.5%) | 15 (10.0%) | 4 (13.8%) |
| Current | 4 (1.9%) | 4 (1.9%) | 3 (2.0%) | 0 (0%) |
| Median BMI, kg/m<sup>2</sup> (Range) | 25.8 [18.0, 40.9] | 25.6 [15.5, 42.6] | 25.4 [15.5, 42.6] | 25.6 [18.3, 34.7] |
| mdNLR mean (SD) | 1.41 (0.67) | 1.48 (0.82) | 1.47 (0.75) | 1.34 (0.77) |
| mdNLR median (range) | 1.31 [0.12, 4.38] | 1.33 [0.26, 7.09] | 1.31 [0.26, 4.50] | 1.19 [0.34, 3.36] |
| CRP Score 1 mean (SD) | 0.02 (1.01) | -0.02 (0.99) | -0.04 (0.97) | 0.47 (1.08) |
| CRP Score 1 median (range) | 0.06 [-2.42, 2.99] | -0.03 [-3.26, 2.94] | -0.03 [-3.26, 2.94] | 0.41 [-1.93, 2.16] |
| CRP Score 2 mean (SD) | 0.04 (1.03) | -0.04 (0.98) | -0.06 (0.95) | 0.36 (1.17) |
| CRP Score 2 median (range) | 0.01 [-3.35, 3.93] | -0.10 [-2.68, 3.10] | -0.11 [-2.68, 3.10] | 0.24 [-2.34, 3.10] |
| CRP Score 3 mean (SD) | -0.06 (0.98) | 0.06 (1.02) | 0.09 (1.03) | 0.32 (0.97) |
| CRP Score 3 median (range) | -0.09 [-2.43, 2.51] | 0.08 [-2.34, 2.95] | 0.10 [-2.34, 2.95] | 0.38 [-2.19, 1.97] |

### Methylation-derived mdNLR index, leukocyte proportions, and lung cancer risk

We observed a 47% increased risk of non-small cell lung cancer (NSCLC) for one standard deviation increase in mdNLR (n = 150 pairs; OR: 1.47 [1.08, 2.02]). However, higher mdNLR values were not statistically associated with overall risk of lung cancer in our study. This association was comparable for NSCLC cases diagnosed within 10 years and beyond 10 years after blood draw. mdNLR was not statistically significantly associated with risk of total lung cancer or small cell lung cancer (SCLC). In addition, immune cell ratios for CD4/CD8, NLR, B-cell/lymphocyte, T-cell/lymphocyte, and monocyte/lymphocyte did not exhibit statistically significant associations with risk of lung cancer, either overall or risk for lung cancer specific subtypes (Table 2).

**Table 2.** Association between for immune cell ratios and methylation-based CRP scores and risk of total lung cancer and NSCLC risk overall and stratified by time to diagnosis, CLUE I/II cohort

|  | All lung cancers <sup>a</sup><br>OR (95% CI) | Non-small Cell<br>lung cancer <sup>b</sup><br>OR (95% CI) |
| --- | --- | --- |
| **mdNLR <sup>c</sup>** | 1.11 (0.89, 1.38) | **1.47 (1.08, 2.02)** |
| time to diagnosis ≤ 10 years | 0.97 (0.69, 1.35) | 1.41 (0.90, 2.20) |
| time to diagnosis > 10 years | 1.22 (0.91, 1.64) | 1.57 (0.98, 2.49) |
| **B cell/Lymphocyte Ratio <sup>d</sup>** | 1.05 (0.84, 1.31) | 0.95 (0.72, 1.26) |
| time to diagnosis ≤ 10 years | 1.07 (0.76, 1.49) | 0.89 (0.57, 1.39) |
| time to diagnosis > 10 years | 1.01 (0.75, 1.36) | 0.98 (0.67, 1.43) |
| **CD4/CD8 Ratio <sup>e</sup>** | 1.04 (0.84, 1.28) | 1.13 (0.86, 1.47) |
| time to diagnosis ≤ 10 years | 0.92 (0.64, 1.30) | 0.86 (0.45, 1.65) |
| time to diagnosis > 10 years | 1.13 (0.83, 1.55) | 1.27 (0.89, 1.82) |
| **T cell/Lymphocyte Ratio <sup>f</sup>** | 1.07 (0.84, 1.37) | 0.94 (0.70, 1.27) |
| time to diagnosis ≤ 10 years | 1.15 (0.76, 1.73) | 0.91 (0.54, 1.54) |
| time to diagnosis > 10 years | 1.05 (0.77, 1.45) | 0.96 (0.65, 1.41) |
| **CRP Score 1 <sup>g</sup>** | **0.76 (0.59, 0.99)** | **0.57 (0.40, 0.81)** |
| time to diagnosis ≤ 10 years | 0.84 (0.56, 1.26) | **0.57 (0.33, 0.97)** |
| time to diagnosis > 10 years | **0.67 (0.47, 0.97)** | **0.53 (0.32, 0.88)** |
| **CRP Score 2 <sup>h</sup>** | **0.77 (0.61, 0.98)** | **0.62 (0.45, 0.84)** |
| time to diagnosis ≤ 10 years | 0.79 (0.54, 1.17) | **0.49 (0.28, 0.85)** |
| time to diagnosis > 10 years | 0.73 (0.53, 1.02) | 0.68 (0.46, 1.00) |
| **CRP Score 3 <sup>i</sup>** | 0.79 (0.58, 1.07) | **0.65 (0.44, 0.95)** |
| time to diagnosis ≤ 10 years | 0.87 (0.53, 1.42) | 0.64 (0.35, 1.18) |
| time to diagnosis > 10 years | 0.73 (0.49, 1.09) | 0.62 (0.36, 1.05) |

<sup>a</sup> 208 cases and 208 controls matched for age at blood draw, sex, smoking status, and model further adjusting for BMI, surrogate variables for batch effects, and a pack-year based smoking methylation score.
<sup>b</sup> 150 cases and 150 controls matched for age at blood draw, sex, smoking status, and model further adjusting for BMI, surrogate variables for batch effects, and a pack-year based smoking methylation score. The matched pairs were kept within each stratum.
<sup>c</sup> OR results reported per 1 unit of SD of mdNLR (0.75).
<sup>d</sup> OR results reported per 1 unit of SD of BL ratio (0.046).
<sup>e</sup> OR results reported per 1 unit of SD of CD4/CD8 ratio (1.45).
<sup>f</sup> OR results reported per 1 unit of SD of TL ratio (0.073).
<sup>g</sup> CpG Score 1 is built using 54 CpG sites.
<sup>h</sup> CpG Score 2 is built using the top 10 highly cell-specific CpG sites.
<sup>i</sup> CpG Score 3 is built using the 10 modestly cell-specific CpG sites.
NOTE: Bold OR and CI values indicate statistical significance.

### Methylation-derived CRP scores and lung cancer risk

CRP Score 1 was built using 54 CpG sites that were previously associated with inflammatory markers, while CRP Score 2 and 3 were each built with a subset of these 54 CpGs that were putative cell-specific or cell type invariant respectively. Using data from a previously published pancreatic cancer dataset (25), all three scores were moderately correlated with log CRP and log IL-6 levels (Table 3). In this nested case-control study, CRP Scores 1, 2, and 3 were not associated with lung cancer risk when taking into account the matching factors and adjusting for BMI and four surrogate variables for batch effects (n = 208 pairs; Score 1 OR: 0.96 [0.77, 1.21]; Score 2 OR: 0.89 [0.71, 1.11]; Score 3 OR: 1.11 [0.89, 1.40]).

**Table 3.** Correlations between methylation-based CRP scores and circulating log-CRP level, log-IL6 level, peripheral blood leukocyte types, BMI, and smoking score residual among controls only.

|  | **Score 1<sup>b</sup>** | **Score 2<sup>c</sup>** | **Score 3<sup>d</sup>** |
| --- | --- | --- | --- |
|  | Spearman (p-value) | Spearman (p-value) | Spearman (p-value) |
| **Pancreatic cancer study (controls only)** |  |  |  |
| Log-CRP level<sup>a</sup> | 0.284 (4.27e-06) | 0.247 (6.96e-05) | 0.297 (1.56e-06) |
| Log-IL6 level<sup>a</sup> | 0.213 (7.53e-04) | 0.158 (1.32e-02) | 0.203 (1.33e-03) |
| **CLUE I/II (controls only)** |  |  |  |
| CD4T | 0.425 (1.65e-10) | 0.359 (1.00e-07) | 0.212 (2.13e-03) |
| CD8T | 0.504 (8.78e-15) | 0.461 (2.34e-12) | 0.390 (5.74e-09) |
| NK | 0.062 (3.70e-01) | 0.175 (1.17e-02) | 0.026 (7.09e-01) |
| B cell | 0.103 (1.38e-01) | 0.069 (3.23e-01) | -0.027 (6.99e-01) |
| Neutrophils | -0.467 (1.19e-12) | -0.422 (2.24e-10) | -0.264 (1.17e-04) |
| Monocytes | -0.292 (1.92e-05) | -0.330 (1.17e-06) | -0.288 (2.38e-05) |
| mdNLR | -0.488 (7.66e-14) | -0.449 (1.01e-11) | -0.281 (3.96e-05) |
| BMI | 0.025 (7.22e-01) | 0.070 (3.12e-01) | -0.063 (3.68e-01) |
| Methylation predicted packyears residual<sup>e</sup> | 0.475 (4.46e-13) | 0.375 (2.43e-08) | 0.632 (1.42e-24) |

<sup>a</sup> Correlations with log-CRP level and log-IL6 level were tested with a pancreatic cancer dataset (25).
<sup>b</sup> CpG Score 1 is built using 54 CpG sites.
<sup>c</sup> CpG Score 2 is built using the top 10 highly cell-specific CpG sites.
<sup>d</sup> CpG Score 3 is built using the 10 modestly cell-specific CpG sites.
<sup>e</sup> We calculated a pack-years methylation score to represent packyears smoked associated methylation alterations. This score correlates with gene expression changes that are affected by smoking.

However, when additionally adjusting for methylation-predicted pack-years, inverse associations with total lung cancer risk were observed for Score 1 (OR: 0.76 [0.59, 0.99]) and Score 2 (OR: 0.77 [0.61, 0.98]). We also observed a 33% decreased risk of lung cancer for one standard deviation increase in CRP Score 1 (OR: 0.67 [0.47, 0.97]) among those with time to diagnosis over 10 years. All three CRP Scores were inversely associated with risk of NSCLC risk after additionally adjusting for methylation-predicted pack-years (n = 150 pairs; Score 1 OR: 0.57 [0.40, 0.81]; Score 2 OR: 0.62 [0.45, 0.84]; Score 3 OR: 0.65 [0.44, 0.95]). In addition, we found statistically significant inverse association between CRP Score 1 and risk of NSCLC among cases diagnosed within 10 years and beyond 10 years, and between Inflammation Score 2 for cases diagnosed within 10 years of blood draw (Table 2).

### Survival analysis

We examined whether the immune cell ratios and CRP Scores were associated with risk of dying of lung cancer among lung cancer cases (Table 4, Figure 1). We observed a 27% increased risk of dying from lung cancer for one standard deviation increase in mdNLR (n = 205 cases deleted 3 cases with person-year = 0 or > 25 years; HR: 1.27 [1.08, 1.50]). A 50% increased risk of dying for one standard deviation of mdNLR was observed for NSCLC cases (n = 149 cases; HR: 1.50 [1.19, 1.89]). Among the NSCLC cases whose mdNLR was from <=10 years before their diagnosis, we found a 78% increased risk of dying for one standard deviation increase in mdNLR (HR: 1.78 [1.17, 2.73]). In comparison, the risk of dying for one standard deviation increase in mdNLR was slightly attenuated to 42% among the NSCLC cases whose mdNLR was from >10 years before their diagnosis (HR: 1.42 [1.03, 1.95]). Immune cell ratios for CD4/CD8, B-cell/lymphocyte, T-cell/lymphocyte, and monocyte/lymphocyte were not associated with lung-cancer specific death, with the exception of a 40% increased risk for one standard deviation increase in B-cell/lymphocyte ratio among the NSCLC cases whose methylation-predicted ratio was from >10 years before diagnosis (HR: 1.40 [1.01, 1.92]). Furthermore, the three CRP Scores were not associated with lung cancer-specific death, except for a 47% decreased risk of dying for one standard deviation increase in CRP Score 1 among the NSCLC cases with BMI ≥ 25 kg/m^2^ (HR: 0.53 [0.28, 1.99]).

**Table 4.**
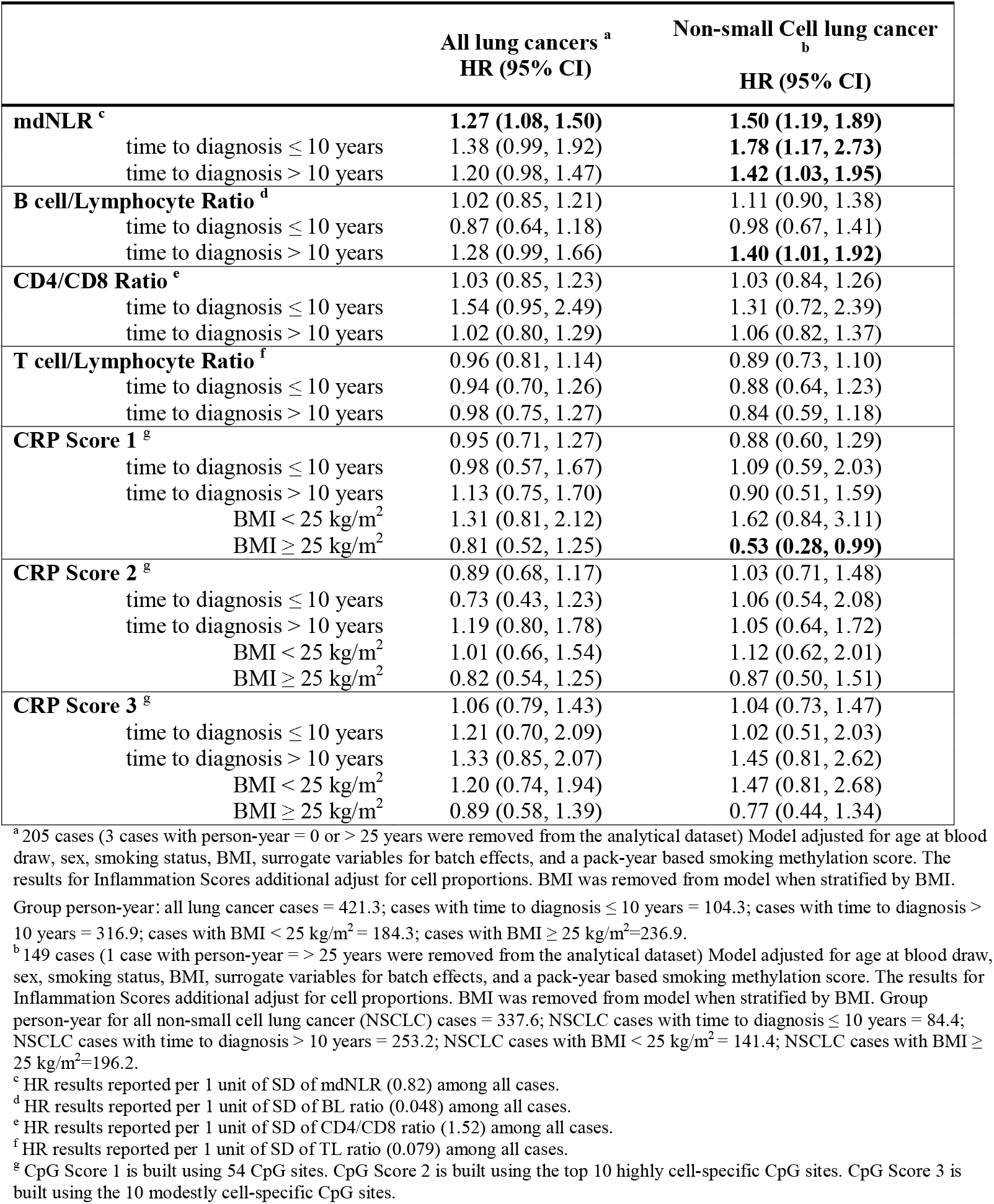
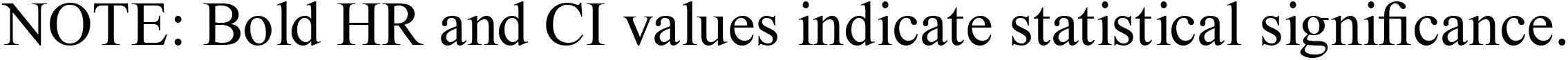
Association between immune cell ratios and methylation-based CRP scores and lung-cancer specific mortality in among lung cancer cases, CLUE I/II cohort

|  | All lung cancers <sup>a</sup><br>HR (95% CI) | Non-small Cell lung cancer <sup>b</sup><br>HR (95% CI) |
| --- | --- | --- |
| **mdNLR <sup>c</sup>** | **1.27 (1.08, 1.50)** | **1.50 (1.19, 1.89)** |
| time to diagnosis ≤ 10 years | 1.38 (0.99, 1.92) | **1.78 (1.17, 2.73)** |
| time to diagnosis $>$ 10 years | 1.20 (0.98, 1.47) | **1.42 (1.03, 1.95)** |
| **B cell/Lymphocyte Ratio <sup>d</sup>** | 1.02 (0.85, 1.21) | 1.11 (0.90, 1.38) |
| time to diagnosis ≤ 10 years | 0.87 (0.64, 1.18) | 0.98 (0.67, 1.41) |
| time to diagnosis $>$ 10 years | 1.28 (0.99, 1.66) | **1.40 (1.01, 1.92)** |
| **CD4/CD8 Ratio <sup>e</sup>** | 1.03 (0.85, 1.23) | 1.03 (0.84, 1.26) |
| time to diagnosis ≤ 10 years | 1.54 (0.95, 2.49) | 1.31 (0.72, 2.39) |
| time to diagnosis $>$ 10 years | 1.02 (0.80, 1.29) | 1.06 (0.82, 1.37) |
| **T cell/Lymphocyte Ratio <sup>f</sup>** | 0.96 (0.81, 1.14) | 0.89 (0.73, 1.10) |
| time to diagnosis ≤ 10 years | 0.94 (0.70, 1.26) | 0.88 (0.64, 1.23) |
| time to diagnosis $>$ 10 years | 0.98 (0.75, 1.27) | 0.84 (0.59, 1.18) |
| **CRP Score 1 <sup>g</sup>** | 0.95 (0.71, 1.27) | 0.88 (0.60, 1.29) |
| time to diagnosis ≤ 10 years | 0.98 (0.57, 1.67) | 1.09 (0.59, 2.03) |
| time to diagnosis $>$ 10 years | 1.13 (0.75, 1.70) | 0.90 (0.51, 1.59) |
| BMI $<$ 25 kg/m<sup>2</sup> | 1.31 (0.81, 2.12) | 1.62 (0.84, 3.11) |
| BMI ≥ 25 kg/m<sup>2</sup> | 0.81 (0.52, 1.25) | **0.53 (0.28, 0.99)** |
| **CRP Score 2 <sup>g</sup>** | 0.89 (0.68, 1.17) | 1.03 (0.71, 1.48) |
| time to diagnosis ≤ 10 years | 0.73 (0.43, 1.23) | 1.06 (0.54, 2.08) |
| time to diagnosis $>$ 10 years | 1.19 (0.80, 1.78) | 1.05 (0.64, 1.72) |
| BMI $<$ 25 kg/m<sup>2</sup> | 1.01 (0.66, 1.54) | 1.12 (0.62, 2.01) |
| BMI ≥ 25 kg/m<sup>2</sup> | 0.82 (0.54, 1.25) | 0.87 (0.50, 1.51) |
| **CRP Score 3 <sup>g</sup>** | 1.06 (0.79, 1.43) | 1.04 (0.73, 1.47) |
| time to diagnosis ≤ 10 years | 1.21 (0.70, 2.09) | 1.02 (0.51, 2.03) |
| time to diagnosis $>$ 10 years | 1.33 (0.85, 2.07) | 1.45 (0.81, 2.62) |
| BMI $<$ 25 kg/m<sup>2</sup> | 1.20 (0.74, 1.94) | 1.47 (0.81, 2.68) |
| BMI ≥ 25 kg/m<sup>2</sup> | 0.89 (0.58, 1.39) | 0.77 (0.44, 1.34) |

<sup>a</sup> 205 cases (3 cases with person-year = 0 or $>$ 25 years were removed from the analytical dataset) Model adjusted for age at blood draw, sex, smoking status, BMI, surrogate variables for batch effects, and a pack-year based smoking methylation score. The results for Inflammation Scores additional adjust for cell proportions. BMI was removed from model when stratified by BMI.
Group person-year: all lung cancer cases = 421.3; cases with time to diagnosis ≤ 10 years = 104.3; cases with time to diagnosis $>$ 10 years = 316.9; cases with BMI $<$ 25 kg/m<sup>2</sup> = 184.3; cases with BMI ≥ 25 kg/m<sup>2</sup> = 236.9.
<sup>b</sup> 149 cases (1 case with person-year = $>$ 25 years were removed from the analytical dataset) Model adjusted for age at blood draw, sex, smoking status, BMI, surrogate variables for batch effects, and a pack-year based smoking methylation score. The results for Inflammation Scores additional adjust for cell proportions. BMI was removed from model when stratified by BMI. Group person-year for all non-small cell lung cancer (NSCLC) cases = 337.6; NSCLC cases with time to diagnosis ≤ 10 years = 84.4; NSCLC cases with time to diagnosis $>$ 10 years = 253.2; NSCLC cases with BMI $<$ 25 kg/m<sup>2</sup> = 141.4; NSCLC cases with BMI ≥ 25 kg/m<sup>2</sup> = 196.2.
<sup>c</sup> HR results reported per 1 unit of SD of mdNLR (0.82) among all cases.
<sup>d</sup> HR results reported per 1 unit of SD of BL ratio (0.048) among all cases.
<sup>e</sup> HR results reported per 1 unit of SD of CD4/CD8 ratio (1.52) among all cases.
<sup>f</sup> HR results reported per 1 unit of SD of TL ratio (0.079) among all cases.
<sup>g</sup> CpG Score 1 is built using 54 CpG sites. CpG Score 2 is built using the top 10 highly cell-specific CpG sites. CpG Score 3 is built using the 10 modestly cell-specific CpG sites.
NOTE: Bold HR and CI values indicate statistical significance.

**Figure 1.**
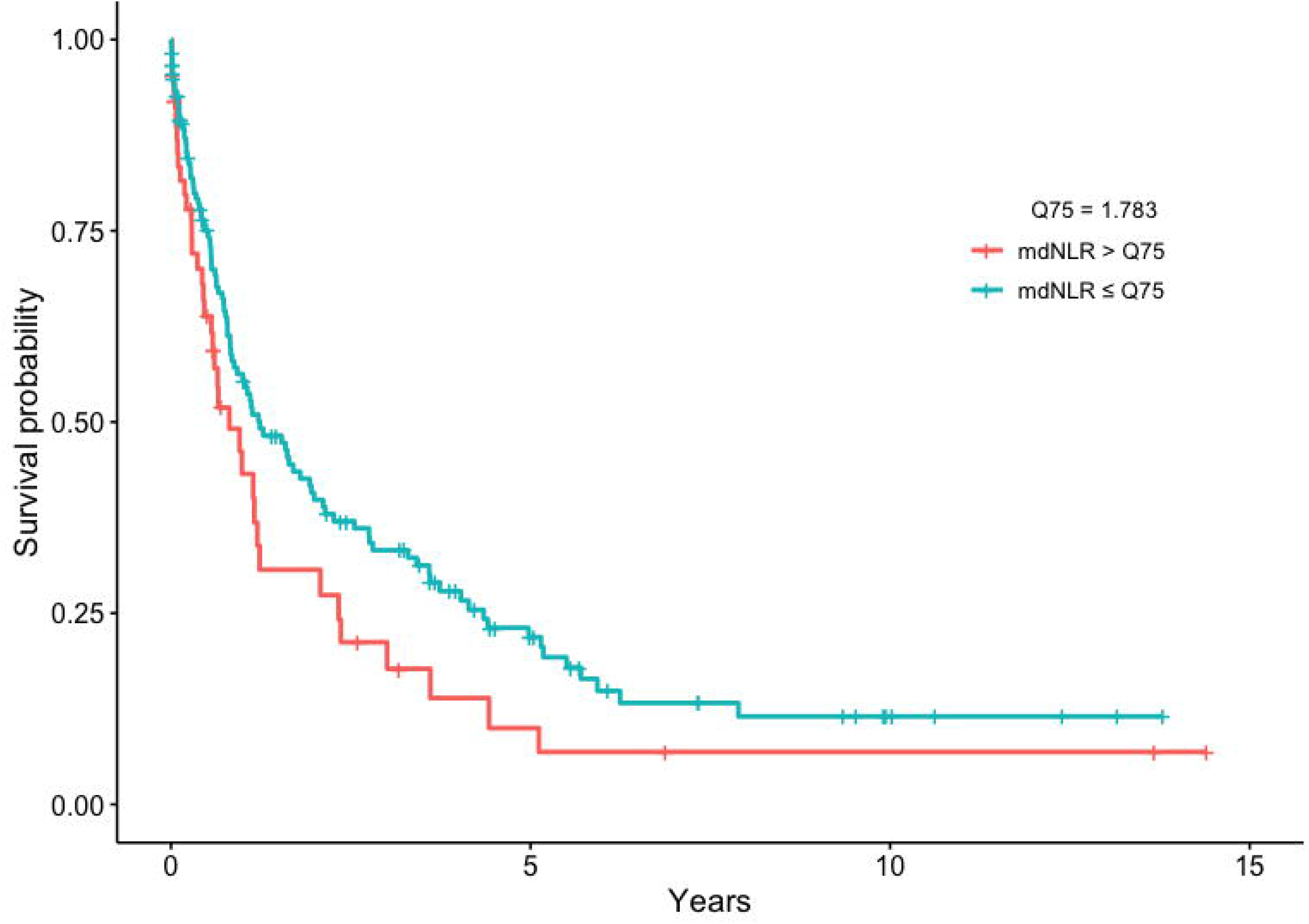
Survival curves for lung cancer specific mortality among lung cancer cases in the mdNLR high and low groups (> or ≤ 75% quartiles). Plot adjusted for age, sex, smoking status, methylation-predicted packyears smoked, BMI, stage, and batch effects.

## Discussion

Our study prospectively assessed predicted immune profiles using DNA methylation markers and examined associations between previously identified DNA methylation markers of inflammation and lung cancer risk and survival. Using pre-diagnostic blood samples of lung cancer cases and controls who participated in the CLUE I/II cohorts [23], pre-diagnosis mdNLR was associated with increased risk of NSCLC, and among cases, with total lung cancer and NSCLC lung cancer specific death. In addition, we built a series of methylation-derived inflammation scores to capture individual systemic inflammatory profiles years before lung cancer diagnosis; these scores were inversely associated with risk of lung cancer, especially for NSCLC after adjusting for methylation-predicted packyears smoked.

Studies on NLR and lung cancer risk and survival typically measure pre-treatment NLR at diagnosis or up to 30 days prior to treatment (26-28). Unlike prior studies, we were able to assess individual systemic inflammation profiles many years prior to diagnosis by using methylation markers of inflammation. Our study is not directly comparable to prior studies since we measured mdNLR using blood samples from subjects with a median of 14 years prior to lung cancer diagnosis. To our knowledge, only one other cohort, the multicenter β-Carotene and Retinol Efficacy Trial (CARET), examined pre-diagnosis mdNLR and lung cancer risk and survival using blood drawn years prior to diagnosis (median 4.7 years) (29, 30). In this study of heavy smokers from CARET, researchers reported a 21% increased risk of lung cancer per one standard deviation increase in mdNLR (OR: 1.21 [1.01, 1.45]), a 30% increased risk of NSCLC for one standard deviation increase in mdNLR (OR: 1.30 [1.03, 1.63], and no association between higher pre-diagnosis mdNLR and risk of developing SCLC (OR: 1.06 [0.77, 1.47]) (29). In contrast, we found no statistically significant association between mdNLR and overall risk of lung cancer in CLUE, but did observe a 47% increased risk of NSCLC for one standard deviation increase in mdNLR (n = 150 pairs; OR: 1.47 [1.08, 2.02]).

CARET researchers recently reported that pre-diagnosis mdNLR is positively associated with increased mortality for SCLC cases, but not for other case types (30). In comparison, we observed a positive association between pre-diagnosis mdNLR and lung cancer-specific and NSCLC-specific mortality. In the case of SCLC, the number of cases was too limited for us to estimate stable associations (N = 29). Taken together, the CLUE and CARET results suggest that a systemic inflammatory profile marked by elevated NLR could indicate a lesser ability to mount a robust immune response to a developing lung cancer and/or a more favorable environment for cancer progression. Differences in findings between the two studies could stem from differences in study populations. The CARET cohort is exclusively heavy smokers, including a subgroup exposed to asbestos. In comparison, our analysis in the CLUE I/II cohorts included never, ever, and current smokers. Furthermore, our study population had a lower mdNLR in the lung cancer cases (mean 1.48 and SD 0.82) than in CARET (mdNLR mean 2.18 and SD 1.46).

We also investigated three CRP Scores that we built from 54 CpG sites that had been strongly associated with CRP in previous studies. We found these CRP Scores to be moderately correlated with log-CRP and log-IL6 in the controls of a previously published pancreatic cancer dataset (25). CRP is a systemic marker of chronic inflammation and has been reported as a risk factor for cancer development (31). Previous studies of pre-diagnostic circulating CRP concentration and lung cancer risk (7 cohorts (8, 9, 17, 32-34) and 3 nested case-control studies (6, 10, 35)) have consistently found a moderate positive association between pre-diagnostic CRP concentrations and lung cancer risk. In our study, CRP Scores were not associated with lung risk when taking into account the matching factors, BMI, and batch effects. However, we observed an inverse association when additionally adjusting for methylation-predicted pack-year. Our results suggest that when strict control of smoking is applied, our CRP Score is likely capturing the unique individual immune response that is not driven by smoking. Furthermore, these results provide preliminary evidence supporting the hypothesis that systemic inflammation not driven by smoking could have a protective effect on individuals. While smoking is by far the most important risk factor for lung cancer, our DNA methylation-based CRP Scores provide the opportunity to examine inflammatory measures not related to smoking that could play a role in modulating cancer risk years prior to diagnosis.

Like other observational studies, our study included a limited number of NSCLC and SCLC cases. The relatively small sample size of SCLC cases (N = 29) impacted our ability to observe associations for this subtype (SCLC comprises about 15% of lung cancer cases in the U.S.). Our study is also limited by a lack of replication dataset and reduced generalizability (study population is mainly Caucasian and with very few cases of never smokers). The CRP Scores we built should be investigated in other populations to ensure that what we observed did not arise due to chance.

## Conclusions

Our study suggests that elevated pre-diagnosis mdNLR and a lower non-smoking-related systemic inflammatory profile before diagnosis are associated with higher cancer risk and poorer lung cancer-specific survival. These relationships were especially evident for NSCLC. As the most common subtype of lung cancer, most NSCLC cases are diagnosed with locally advanced or metastatic disease. Our prospective results support future evaluation of whether DNA methylation-based inflammatory measures could enhance lung cancer risk stratification to improve targeted lung cancer screening.

## Methods

### Study Population

This nested case-control study selected cases and controls from individuals who participated and provided blood in both CLUE I and CLUE II (24). The CLUE I cohort was developed to identify serologic precursors of cancer and was conducted in Washington County, Maryland, in the fall of 1974. A blood sample was collected from 25,620 volunteers at the time of participation (36, 37). The CLUE II cohort was conducted from May through October 1989. During this time, all participants donated a blood sample which was collected in tubes containing heparin from 32,894 individuals and kept chilled until centrifuged, aliquotted into plasma, erythrocytes, and buffy coat and frozen at 70° C (38). In CLUE II, the baseline for this study, health information was collected at the time of blood draw, including attained education, cigarette smoking status, cigarette smoking dose, cigar/pipe smoking status, and self-reported weight and height.

Incident lung cancer cases were ascertained from linkage to the Washington Co. cancer registry (before 1992 to the present) and the Maryland Cancer Registry (since 1992 when it began to the present). We ascertained 241 incident lung cancer cases who participated in CLUE I and were diagnosed after the day of blood draw in CLUE II through January 2018. Cases were characterized with respect to histology. We used incidence density sampling to select one control matched to each case on age, sex, and smoking intensity. The Institutional Review Board at the Johns Hopkins Bloomberg School of Public Health and the Tufts University Health Sciences Campus Institutional Review Board approved this study.

### DNA methylation measurements

Extracted DNA was bisulfite-treated using the EZ DNA Methylation Kit (Zymo) and DNA methylation was measured with the 850K Illumina Infinium MethylationEPIC BeadChip Arrays (Illumina, Inc, CA, USA). All samples and all array experiments were performed blinded to case control status. Details on DNA methylation measurements, data preprocessing processing and quality control assessment/screening are provided in the Supplementary Methods. The 850K methylation microarray has been validated from a biological and technical standpoint. Reproducibility of results from 850K Illumina array has been previously shown to be very high (r=0.997) (39). DNA volume and quality was sufficient for 208 of the cases and 222 controls for a total of 208 matched pairs.

### Estimation of peripheral blood leukocyte composition

Peripheral blood leukocyte subtypes proportions, including CD4+ T cells (CD4T), CD8+ T cells (CD8T), B cells (Bcell), neutrophils (Neu), natural killer cells (NK), and monocytes (Mono) were estimated using the “estimateCellCounts2” function in the FlowSorted.Blood.EPIC Bioconductor package (40), which is based on previously published reference-based cell mixture deconvolution algorithm with reference library selection conducted using the IDOL methodology (41).

### Methylation-Derived Neutrophil Lymphocyte Ratio (mdNLR)

The peripheral blood neutrophil–to-lymphocyte ratio (NLR) is a cytological marker of both inflammation and poor outcomes in cancer patients (42-46). We used a DNA methylation-derived NLR (mdNLR) index to predict the common clinical NLR parameter using a previously described approach (7). This index is based on normal isolated leukocyte reference DNA methylation libraries and established reference-based cell-mixture deconvolution algorithms (7, 47).

### Inflammation-associated CpG score

We selected 54 CpG sites that have been strongly associated with inflammatory measures to build a CRP Score (48, 49). Ligthart and colleagues identified 58 CpGs (48) that were associated with serum C-reactive protein (CRP) levels (listed in their Table 2) using 450K DNA methylation data. 45 of these 58 CpG sites were validated to have same direction of protein-methylation associations by Myte et al (49). We used 54 of the 58 CpG sites reported by Ligthart et al. in this study since the remain 4 CpG sites were not contained on the 850K array we used to measure methylation. To compute the CRP Score, we multiplied the beta value at each selected CpG sites with the effect size estimates reported by Ligthart et al. These estimated beta coefficients represented the change in DNA methylation per one unit increase in log CRP. In the CRP Score formula, we weighted the beta coefficients estimated by Ligthart et al. with their corresponding standard errors.

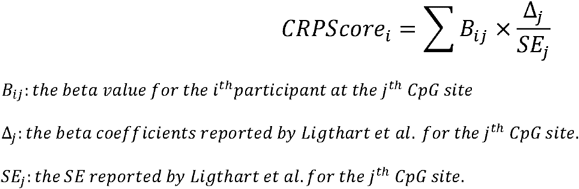

We named this score CRP Score 1. Since most of the estimated beta coefficients are negative, CRP Score 1 ranged between -0.059 and -0.026. A score closer to zero indicated higher CRP levels. Based on CRP Score 1, we computed two additional CRP Scores, one cell (leukocyte)-type invariant (CRP Score 2) and one cell-specific (CRP Score 3). Among the 54 inflammation (CRP)-associated CpGs we identified putative cell-type invariant and cell-specific CpGs by conducting ANOVA analysis using the data set described in Salas & Koestler et al. (40) and publicly available on the Gene Expression Ombinbus (GSE110555). The data set used for this ANOVA analysis consisted of EPIC methylation data profiled in purified leukocyte cell population isolated from different healthy adults. Specifically, methylation signatures were available for CD4T, CD8T, NK, Bcell, Monocytes, and Neutrophils. One-way ANOVA models were fit independently to each of the 54 CRP-associated CpGs treating methylation as the dependent variable and cell type as the independent variable. We tested the null hypothesis that the mean methylation beta-value is the same across the cell types. The F-statistic, corresponding p-value, and maximum absolute pairwise difference in the mean methylation beta value across cell types were calculated for each of the 54 CpGs. We then selected subgroups of CpG sites that had the top 10 smallest or top 10 largest F-statistic value to build two additional CRP Scores. CRP Score 2 consists of putative cell-specific CpGs with high F-statistics, e.g., those exhibiting a difference in mean methylation beta-values between at least two of the six cell types. CRP Score 3 is made of cell type invariant CpGs with low F-statistics, e.g., CpGs for which there did not appear to be a substantial difference in mean methylation beta-values across the normal six leukocyte subtypes. Score 2 ranged between -0.0002 and 0.0046 while Score 3 ranged between -0.025 and -0.016. In the subsequent statistical analyses, we used a standardized version of CRP Score 1, 2, and 3 (mean = 0, sd = 1) in all regressions for easier interpretation of results.

### Statistical analyses

All statistical analyses were performed in R (version 3.5.1). Immune cell ratios (e.g., CD4/CD8, neutrophil/lymphocyte, B cell/lymphocyte, T cell/lymphocyte) were calculated for each sample by taking the ratio of its predicted cell proportions described above. We used a pancreatic cancer dataset to estimate the Spearman’s rank correlation and Pearson correlation between estimated values of CRP Scores 1, 2, and 3 with the log CRP and log IL-6 levels (25).

To account for the 1:1 case/control matching structure in our data, a series of multivariable conditional multivariable logistic regression models were used to examine the association between DNA methylation-based inflammatory measures (CRP Scores 1-3 and continuous mdNLR) and lung cancer risk. Models were fit with age, sex, and smoking status (never, former, current) as matching factors, and were adjusted for potential confounding factors, including body mass index (BMI), batch effect, and previously described methylation-predicted pack-years smoked (50). We repeated these analyses by lung cancer histology (NSCLC, SCLC), length of time between blood draw and diagnosis (<=10, >10 years), and BMI (<25, ≥ 25 kg/m^2^). Among the lung cancer cases, we examined the association between these same pre-diagnostic DNA methylation-based inflammatory measures (CRP Scores 1-3 and continuous mdNLR) and risk of lung cancer specific death using a series of multivariable Cox proportional hazard regression adjusting for age, gender, smoking status, BMI, stage (three strata: stage 1 & 2, stage 3 & 4, and missing), cell proportion, batch effects, and methylation-predicted pack-years smoked.

## Data Availability

The datasets generated during the current study are available from the corresponding author on reasonable request and will be deposited into dbGaP by publication.

## Declarations

### Ethics approval and consent to participate

The Institutional Review Board at the Johns Hopkins Bloomberg School of Public Health and the Tufts University Health Sciences Campus Institutional Review Board approved this study.

### Consent for publication

NA

## Authors’ contributions

DSM., KTK and EAP designed the study, obtained funding and acquisition of data. JL assisted with preparation of dataset. DSM supervised all research activities. MR and NZ conducted the statistical analyses. NZ drafted the manuscript. DSM., EAP, KTK, and DCK interpreted the data and provided critical revisions of the manuscript. All authors read and approved the final version of the manuscript.

## Acknowledgements

Cancer incidence data were provided by the Maryland Cancer Registry, Center for Cancer Surveillance and Control, Maryland Department of Health, 201 W. Preston Street, Room 400, Baltimore, MD 21201. We acknowledge the State of Maryland, the Maryland Cigarette Restitution Fund, and the National Program of Cancer Registries of the Centers for Disease Control and Prevention for the funds that helped support the availability of the cancer registry data.

