## Supplemental Methods for "Methylation-derived Inflammatory Measures and Lung Cancer Risk and Survival"

**Supplementary Methods**

**Study population – CLUE I/II cohort**

All cases and controls in this study were selected from individuals that participated and provided blood in both CLUE I and CLUE II [1-3]. The CLUE I cohort was developed to identify serologic precursors of cancer and was conducted in Washington County, Maryland, in the fall of 1974. The CLUE II cohort was conducted from May through October 1989. In both CLUE I and II, participants were recruited using mobile office trailers and blood was drawn into 20 mL heparinized Vacutainers (Becton-Dickinson, Rutherford, NJ). All samples were kept at 4°C until the serum (CLUE I) or plasma (CLUE II) was separated, usually within 2–6 h, and divided into aliquots of serum (CLUE I) or plasma, buffy coat, and red blood cells (CLUE II). Comparisons with published figures from the 1980 Census indicated that approximately 30% of adult residents had participated in CLUE II. Women, Caucasians and the better-educated had higher participation rates, as did the age group 45 to 70 years. The CLUE cohort is community-based with reasonable representation of the general county population, with substantial heterogeneity of risk factors for disease outcomes. Participants were sent follow-up questionnaires in 1996, 1998, 2000, and 2007. Participants were sent newsletters periodically with forwarding addresses requested from the United States Postal Service.

**Sample Selection**

We selected all men and women in whom lung cancers were diagnosed after the date of blood draw (CLUE II) through January 2018, who did not have a prior cancer diagnosis (except for nonmelanoma skin cancer or cervix in situ), and who participated in CLUE I. All 241 lung cancer cases (ICD 9 162 and ICD10 C34) were confirmed by pathology report. Controls was selected from among CLUE I/II participants who did not have a diagnosis of cancer and were known to be alive at the time the case was diagnosed. One control was matched to each case on the following factors: age (±1 year), sex, race, cigarette smoking status and number of cigarettes smoked, cigar/pipe smoking status, and date of blood draw (±2 weeks).

Due to lack of DNA, 8 of the 241 incident cases were removed from the dataset before matching. We sent a total of 418 samples (233 cases and 233 controls) to University of Minnesota Genomics Center for processing. When controls had poor DNA quality, we attempted to identify new controls to replace them. During DNA methylation profiling, an additional 19 subjects were removed due to insufficient amount of DNA and 16 samples were removed due to low quality DNA. In the end, 383 samples from 430 subjects (208 cases and 222 controls) passed data quality control. Supplemental Figure 1 documented the selection of study subjects and samples for the final analysis dataset.

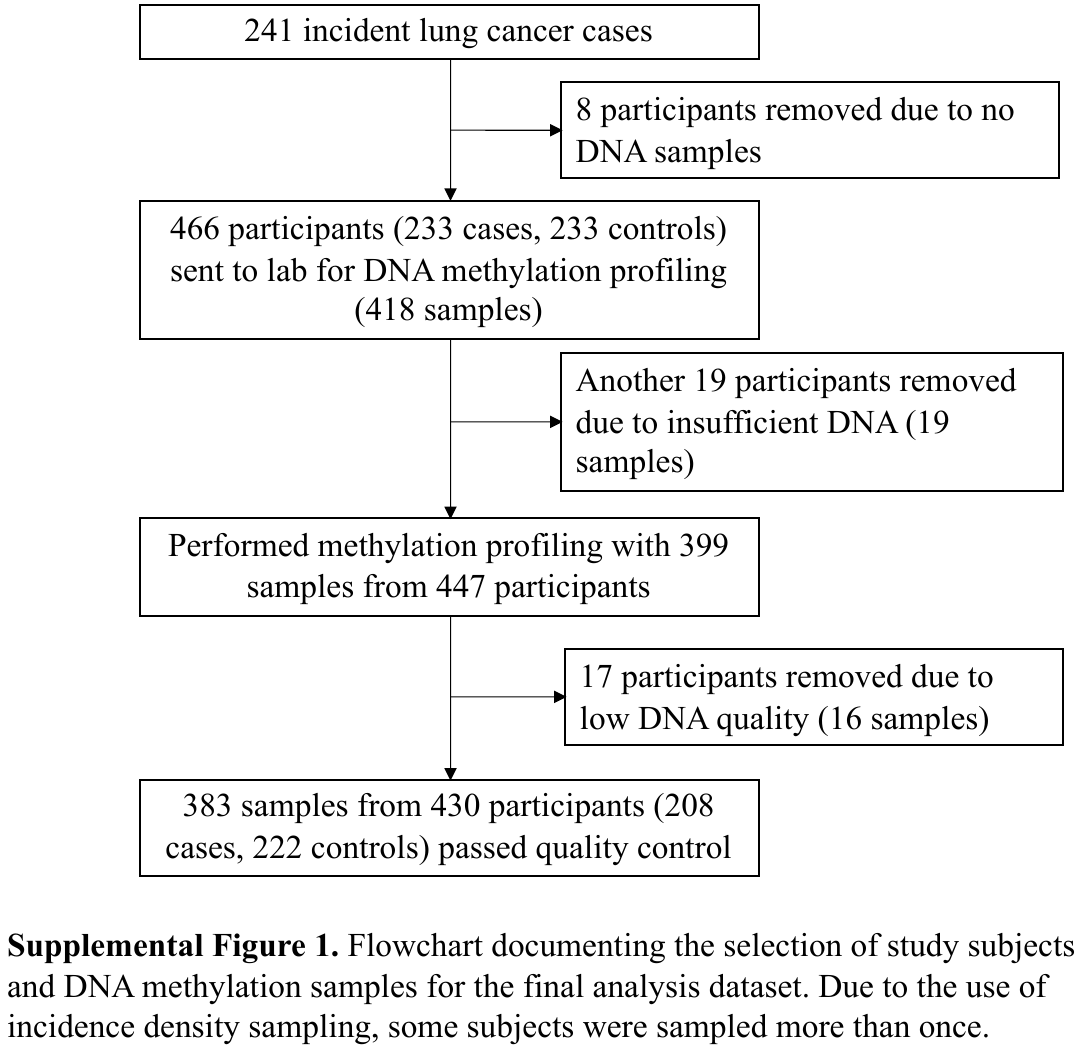

**DNA methylation assay**

Samples of buffy coat were retrieved from freezers for DNA extraction using Qiagen kits. Samples were quantified by PicoGreen and required volume of DNA was plated in 96-well plates. Bisulfite conversion and array processing DNA (previously extracted for genotyping studies in these cohorts) was sent to University of Minnesota Genomics Center for processing. Bisulfite conversion was performed using the EZ DNA Methylation Kit (Zymo). Post-conversion QC was conducted to determine the amount of covered DNA and the completeness of conversion. The samples were processed using the recently released Illumina MethylationEPIC BeadArrays, which interrogates >850,000 CpG sites per sample. Laboratory personnel were blinded to case-control status when performing all array experiments. The 850K methylation microarray has been validated from a biological and technical standpoint. Reproducibility of results from the 850K Illumina array technology has been previously shown to be very high (r=0.997) [4].

**Data processing**

All methylation data preprocessing and normalization were performed using the Bioconductor packages. The raw IDAT files from methylation array were read in using the minfi Bioconductor package [5, 6]. Within-array correction for background fluorescence and dye-biases were performed using the Noob methodology via the function “preprocessNoob” in the minfi Bioconductor package [7]. The QCinfo function in ENmix Bioconductor [8] package was then used to identify and remove poor quality samples and probes [8]. Samples were excluded if: 1) more than 5% of probes had quality issues as addressed using the detection p-value, 2) the bisulfite conversion intensity was lower than 3 standard deviations from the mean, or 3) the mean average intensity and/or the mean average beta values were more than 3 times IQR from the upper quartile or less than 3 time IQR from the lower quartile of the average intensity values or beta value across the samples. In addition, we excluded probes that had detection p-values exceeding 1x10 -6 (compared to the negative background probes) in more than 5% of the samples. After sample- and probe- level quality control, we corrected the type II probe bias to make the methylation distribution of type II feature comparable to the distribution of type I feature using the beta mixture quantile dilation intra-sample normalization method[9], implemented using “BMIQ” function in the wateRmelon Bioconductor package [10]. Principal components analysis (PCA) was performed on the BMIQ-adjusted values and the top K principal components (K determined using a previously described random matrix theory approach [11]) to detect whether the microarray dataset had the batch effect. Then *ctrlsva* function in ENmix Bioconductor packages [12, 13] was used to estimate the surrogate variables of batch effects [14]. The estimated surrogate variables were used in downstream analyses to remove the batch effects and other unwanted experimental confounders.

**Pack-years methylation score**

The pack-years methylation score is calculated to represent packyears smoked-associated methylation alterations [15]. This packyears methylation score was first developed to predict smoking pack-years using smoking ‘signatures’ reported by large-scale epigenome-wide association meta-analyses [15]. This score correlates with gene expression changes that are affected by smoking, and can be utilized in lieu of self-reported smoking data [15].
